# Automated Expert-level Scleral Spur Detection and Quantitative Biometric Analysis on the ANTERION Anterior Segment OCT System

**DOI:** 10.1101/2022.08.23.22279135

**Authors:** Kyle Bolo, Galo Apolo, Anmol A. Pardeshi, Michael Chiang, Bruce Burkemper, Xiaobin Xie, Alex S. Huang, Martin Simonovsky, Benjamin Y. Xu

## Abstract

**Aim:** To perform an independent validation of deep learning (DL) algorithms for automated scleral spur detection and measurement of scleral spur-based biometric parameters in anterior segment optical coherence tomography (AS-OCT) images.

**Methods:** Patients receiving routine eye care underwent AS-OCT imaging using the ANTERION OCT system (Heidelberg Engineering, Heidelberg, Germany). Scleral spur locations were marked by three human graders (Reference, Expert, and Novice) and predicted using DL algorithms developed by Heidelberg Engineering that prioritize a false positive rate <4% (FPR4) or true positive rate >95% (TPR95). Performance of human graders and DL algorithms were evaluated based on agreement of scleral spur locations and biometric measurements with the Reference Grader.

**Results:** 1,308 AS-OCT images were obtained from 117 participants. Median differences in scleral spur locations from reference locations were significantly smaller (p<0.001) for the FPR4 (52.6±48.6μm) and TPR95 (55.5±50.6μm) algorithms compared to the Expert (61.1±65.7μm) and Novice (79.4±74.9μm) Graders. Inter-grader reproducibility of biometric measurements was excellent overall for all four (intraclass correlation coefficient [ICC] range 0.918-0.997). Inter-grader reproducibility of the Expert Grader [0.567-0.965] and DL algorithms [0.746-0.979] exceeded that of the Novice Grader [0.146-0.929] for images with narrow angles, defined as angle opening distance 500μm from the scleral spur (AOD500) <150μm.

**Conclusions:** DL algorithms on the ANTERION approximate expert-level measurement of scleral spur-based biometric parameters in an independent patient population. These algorithms could enhance clinical utility of AS-OCT imaging, especially for evaluating patients with angle closure and performing intraocular lens (IOL) calculations.

**KEY MESSAGES:** 

**What is already known on this topic:** Deep learning (DL) algorithms can detect scleral spur locations in AS-OCT images with expert-level performance; however, there is sparse information about the accuracy of AS-OCT measurements associated with these predicted scleral spur locations.

**What this study adds:** DL algorithms on the ANTERION OCT system (Heidelberg Engineering, Heidelberg, Germany) approximate expert-level detection of the scleral spur and measurement of anterior segment biometric parameters in a real-world clinical cohort. Performance of the algorithms generally exceeds that of a novice grader.

**How this study might affect research, practice or policy:** The automation of scleral spur detection and quantitative biometric analysis overcomes the time-and expertise-dependent nature of AS-OCT imaging in the clinical setting. This technology provides clinicians with convenient access to data that could enhance care of patients with angle closure disease or patients receiving intraocular lens implantation.

## INTRODUCTION

The biometric properties of the anterior segment and its anatomical structures play an important role in the clinical care of patients with a range of ocular conditions. Specifically, anterior segment biometrics play an important role in the pathogenesis of primary angle closure disease (PACD), in which aqueous humor outflow is impaired by apposition of the iris and trabecular meshwork, and closure of the anterior chamber angle (ACA).^1–3^ This process leads to primary angle closure glaucoma (PACG), a major cause of visual morbidity worldwide that currently affects more than 20 million people.^4,5^ In addition, the surgical treatment of eyes with cataract and high refractive error benefits from accurate biometric measurements when calculating the power and size of intraocular lenses (IOLs). Incorrect lens power leads to poor visual outcomes, and incorrect lens sizing can lead to harmful complications such as hyphema, uveitis, glaucomatous optic neuropathy, or corneal decompensation.^6,7^

There is growing evidence that supports the clinical utility of anterior segment optical coherent tomography (AS-OCT) for measuring anterior segment biometrics, many of which are based on scleral spur location. For example, angle opening distance (AOD) and trabecular iris space area (TISA), may find expanded roles in predicting progression of PACD and response to treatment with laser peripheral iridotomy (LPI).^3,8,9^ Quantitative OCT-based methods could complement gonioscopy, which remains the current standard for assessing the ACA despite being subjective, qualitative, variably reproducible, and weakly correlated with AS-OCT measurements of angle width.^10–16^ In IOL selection, biometric parameters, including corneal curvature, anterior chamber depth (ACD), and lens thickness (LT), are measured using optical or ultrasound methods and factored into modern IOL calculators.^17^ Anterior chamber width (ACW), also referred to as white-to-white (WTW) distance, is important for sizing anterior chamber and phakic IOLs.^18–21^ Biometric parameters based on scleral spur location, such as lens vault (LV) and anterior chamber width (ACW), are potentially useful in IOL selection, but are difficult to measure and therefore rarely used in routine clinical practice.^22,23^

Full biometric analysis of AS-OCT images on commercial devices currently requires specialized software and manual marking of scleral spurs, which is expertise-dependent and time-consuming, thereby presenting a barrier to widespread implementation.^24,25^ Prior studies have established the accuracy of scleral spur detection automated using deep learning (DL), a form of artificial intelligence.^26,27^ In this study, we investigate if biometric measurements associated with scleral spur locations predicted by DL algorithms on the Heidelberg ANTERION version 1.4 swept-source OCT system (Heidelberg Engineering GmbH, Heidelberg, Germany) approximate inter-expert reproducibility in an independent patient population and clinical environment.

## METHODS

The study was approved by the University of Southern California (USC) Institutional Review Board. All study procedures adhered to the recommendations of the Declaration of Helsinki. Written informed consent was obtained from all participants.

### Scleral spur detection algorithm

DL algorithms to automate scleral spur detection were developed and tested internally by Heidelberg Engineering (Heidelberg, Germany) prior to this study. While these algorithms are proprietary, some information was provided by Heidelberg Engineering about their development. In brief, a set of 4,798 ANTERION AS-OCT images from one or both eyes of 360 patients were evaluated by an expert ophthalmologist to identify scleral spur locations. These images were divided into non-overlapping training (3,810 images; 80%) and test (979 images; 20%) datasets. The training dataset was used to train a convolutional neural network (CNN) based on the M2U-Net architecture that predicts scleral spur location within a predefined region of interest (ROI).^28^ The ROI is a 256×256 pixel area around the ACA determined heuristically based on the posterior boundary of the cornea and the anterior boundary of the iris as defined by the ANTERION’s segmentation algorithms. Reference scleral spur locations were transformed into reference heatmaps containing a Gaussian function with standard deviation of 10 pixels centered on the reference location. Data augmentation, including affine deformations, noising, and blurring was used to increase the robustness of the CNN. The subpixel-refined position and intensity of the strongest peak in the predicted heatmap were used to estimate the position and confidence (level of certainty ranging from 0 to 1) of the scleral spur. The test dataset was used to select two operating points along the receiver operating characteristic (ROC) curve (Supplementary Figure 1) for further analysis, one more conservative to limit the false positive rate (FPR; scleral spur marked by the algorithm but not the ophthalmologist) below 4% (FPR4 algorithm) and the other more aggressive to ensure a true positive rate (TPR; scleral spur marked by the algorithm and ophthalmologist) above 95% (TPR95 algorithm).

### Acquisition and analysis of validation dataset

Participants 18 years of age and older undergoing routine eye examinations were consecutively recruited from eye care facilities of the Roski Eye Institute at the University of Southern California (USC) and Doheny Eye Institute at the University of California Los Angeles (UCLA). Recruitment occurred from March 2021 to August 2021. Exclusion criteria included corneal opacities that precluded AS-OCT imaging and prior history of ocular trauma.

Anterior segment OCT imaging was performed using the ANTERION and Metrics Application. All images were obtained by trained technicians following a standardized imaging protocol. Imaging of both eyes was performed in the seated position prior to pupillary dilation in a dark room under standardized lighting conditions (<0.01 lux) at the imaging plane. Participants were instructed to maintain fixation on the internal fixation target with their eyelids open without retraction by the technician.

The scleral spur was identified as the inward projection at the junction of the sclera and cornea.^29^ Scleral spur locations in all six B-scans (separated by 30 degrees, creating 12 angle sectors at 0, 30, 60, 90, 120, 150, 180, 210, 240, 270, 300, and 330 degrees) were marked by three human graders: 1) an expert trained grader (AAP; Reference Grader) with experience marking over 40,000 scleral spurs after a 5-hour training period of marking 500 scleral spurs under the supervision of two glaucoma specialists; 2) an expert glaucoma specialist with experience marking over 10,000 scleral spurs (BYX; Expert Grader); 3) a novice trained grader (AS; Novice Grader) with experience marking fewer than 100 scleral spurs. The Reference Grader previously demonstrated low intra-grader variability in scleral spur locations and AS-OCT measurements of biometric parameters.^27,30^ Scleral spur locations were also predicted by the FPR4 and TPR95 algorithms.

The anterior and posterior boundaries of the cornea and lens, and the anterior boundary of the iris were computed automatically by the ANTERION’s segmentation algorithm. The Reference Grader made minor segmentation adjustments in fewer than 15 images (1.1% of total). After the scleral spurs were marked, eight scleral spur-based biometric parameters were measured in an automated fashion by the ANTERION software: angle opening distance (AOD), trabecular iris space area (TISA) and scleral spur angle (SSA) at 500 and 750 µm from the scleral spur, anterior chamber width (ACW), and lens vault (LV). AOD500/750 were defined as the perpendicular distance from the TM at 500 or 750 µm anterior to the scleral spur to the anterior iris surface. TISA500/750 were defined as the area bounded anteriorly by AOD500/750; posteriorly by a line drawn from the scleral spur perpendicular to the plane of the inner scleral wall to the opposing iris; superiorly by the inner corneoscleral wall; and inferiorly by the iris surface. SSA500/750 were defined as the angles formed by lines originating at the scleral spur and terminating at the TM or anterior iris surface 500 or 750 µm anterior to the scleral spur. ACW was defined as the distance between scleral spurs. LV was defined as the perpendicular distance from the apex of the anterior lens surface to a line between scleral spurs.

A subset of images was classified as having narrow angles, defined as an AOD500 measurement below 150 µm by the Reference Grader. This threshold was chosen to define narrow angles due to its high sensitivity and specificity for detecting gonioscopic angle closure.^31^ Narrow angles was not defined based on gonioscopy for several reasons: 1) the majority of patients were not glaucoma patients and therefore did not receive gonioscopy; 2) there is intergrader variability in the detection of gonioscopic angle closure; 3) angle widths associated with gonioscopic angle closure vary significantly by quadrant.^10,32 31^

Images with borderline or poor interpretability due to eyelid and other imaging artifacts were included in the analysis so that false negative rates (FNRs) and false positive rates (FPRs) could be calculated for the Expert and Novice Graders and both DL algorithms. In addition, human graders were not provided specific instruction about what constituted a gradable scleral spur; the decision to grade an image was left to the discretion of each grader. A reference false negative (FN_ref_) was defined as a scleral spur identified by the Reference Grader but not by another grader or algorithm. A reference false positive (FP_ref_) was defined as a scleral spur marked by another grader or algorithm but not by the Reference Grader. A consensus false negative (FN_con_) was defined as a scleral spur marked by all three human graders but not by a DL algorithm. A consensus false positive (FP_con_) was defined as a scleral spur marked by a DL algorithm but not any of the three human graders.

### Statistical analysis

Scleral spur location differences were calculated as the Euclidean distance between scleral spur locations by the Reference Grader and second human grader or DL algorithm. Normality testing was performed on scleral spur location differences using the Kolmogorov-Smirnov test. Medians and interquartile ranges (IQRs) were calculated based on non-normality of the data. Scleral spur location differences were grouped by grader or algorithm and compared using the Kruskal-Wallis test. Pairwise comparisons of scleral spur location differences between groups (six comparisons in total) were performed using the post-hoc Dunn’s test adjusted for multiple comparisons at a significance level of 0.05. Intraclass correlation coefficients (ICCs) were calculated for each biometric parameter measured in all AS-OCT images to assess the inter-grader agreement between the Reference Grader and a second human grader (Expert or Novice) or DL algorithm (FPR4 and TPR95). ICCs were also calculated for each biometric parameter measured in a single sector (superior or temporal) of only one eye per participant to eliminate intra-eye and intra-participant correlations. Bland-Altman plots were generated for AOD500 to assess inter-grader agreement across the entire range of angle widths. All analyses were performed using the R statistical package (version 4.0.3) at a significance level of 0.05.

## RESULTS

In total, 1,308 AS-OCT images were obtained from 117 participants. Mean age was 52.1 ± 17.6 years with 59 males (50.4%) and 58 females (49.6%). Among all participants, 50 (N = 42.7%) were Caucasian, 32 (27.4%) were Hispanic, 21 (17.9%) were Asian, and 7 (6.0%) were Black, and 7 (6.0%) had unknown race/ethnicity.

In total, the Reference Grader marked 1,504 spurs, the Expert Grader marked 1,726 spurs, the Novice Grader marked 1,622 spurs, the FPR4 algorithm marked 1,459 spurs, and the TPR95 algorithm marked 1,722 spurs. Distributions of scleral spur location differences compared to the Reference Grader varied by grader or algorithm (Figures 1 and 2). Median and interquartile range (IQR) of scleral spur location differences were 61.1 ± 65.7 μm for the Expert Grader, 79.4 ± 74.9 μm for the Novice Grader, 52.6 ± 48.6 μm for the FPR4 algorithm, and 55.5 ± 50.6 μm for the TPR95 algorithm. There were significant differences (p < 0.001) among the 4 groups of scleral spur location differences. Pairwise comparisons demonstrated a non-significant difference in scleral spur location differences between the DL algorithms (p = 0.33) and significant differences between all other pairs of graders and algorithms (p < 0.001).

**Figure 1.**
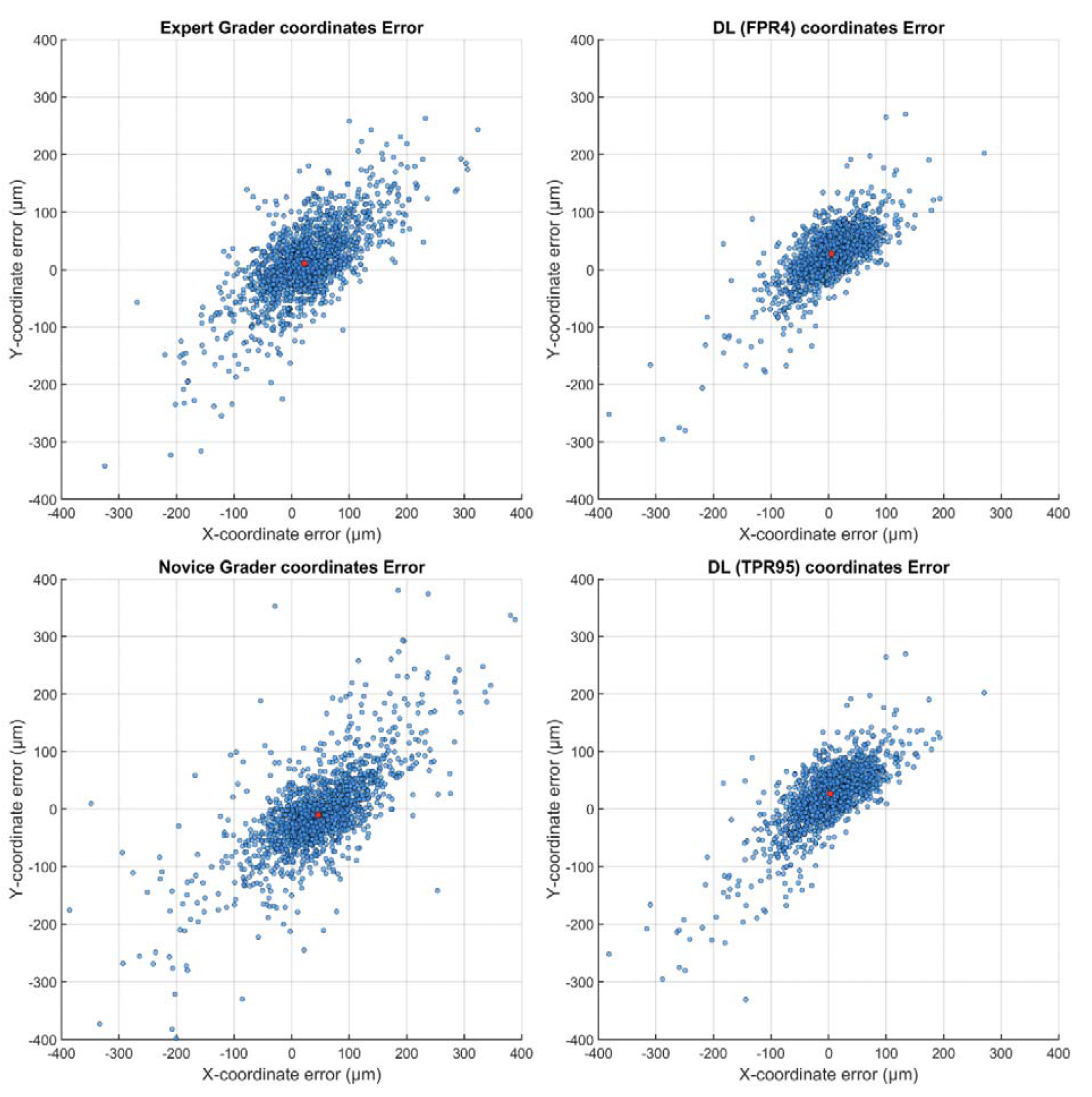
Human-human and human-machine differences in scleral spur locations. Scatter plots showing X-and Y-coordinate errors in comparison to the Reference Grader for the Expert Grader (top left), Novice Grader (bottom left), FPR4 algorithm (top right) and TPR95 algorithm (bottom right). Red dots indicate median X-and Y-coordinate differences.

**Figure 2.**
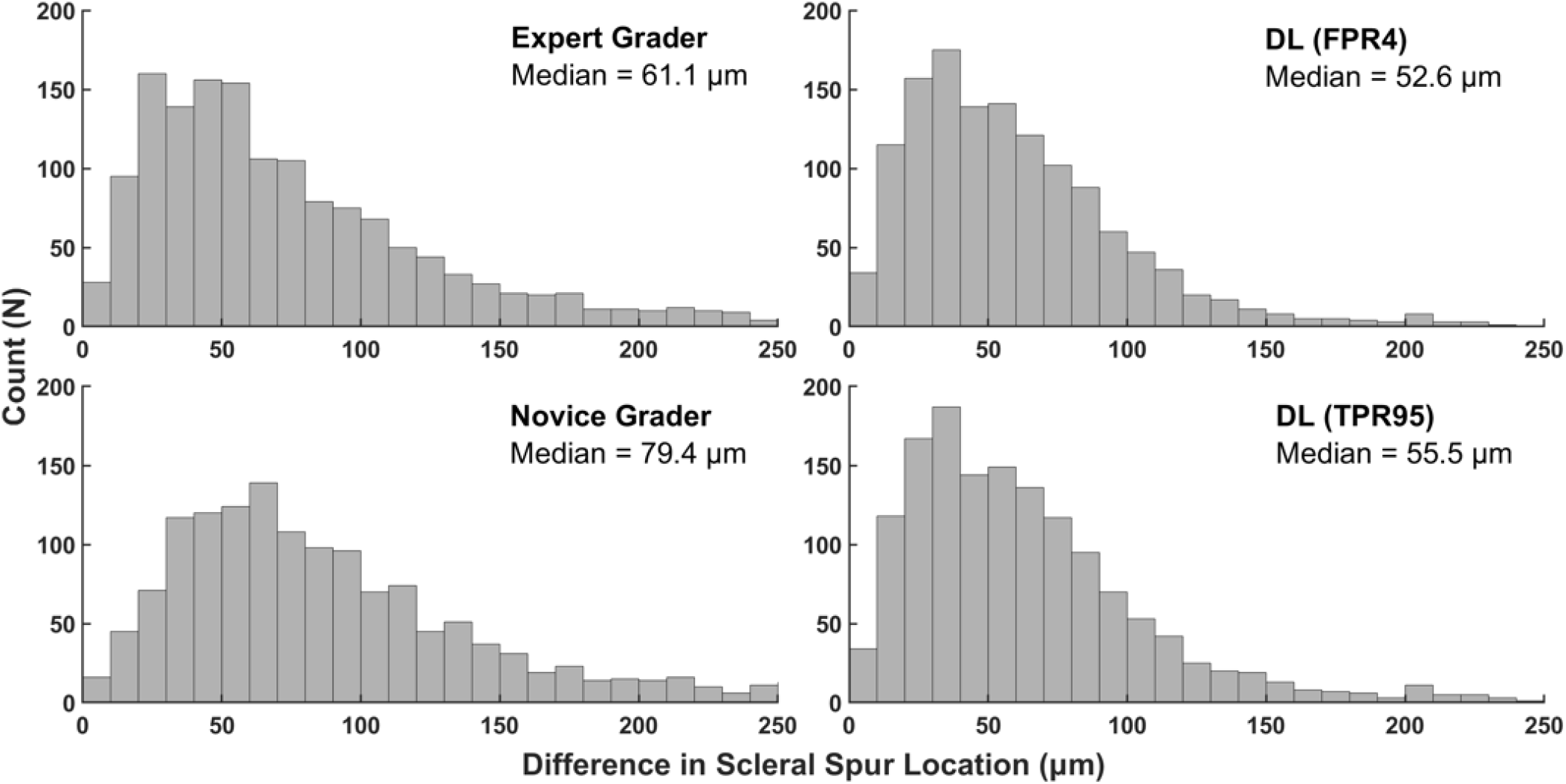
Human-human and human-machine differences in scleral spur locations. Histograms showing the Euclidean distance between scleral spur locations by the Reference Grader and Expert Grader (top left), Novice Grader (bottom left), FPR4 algorithm (top right) and TPR95 algorithm (bottom right).

There was a wide range of angle widths (mean 0.41 ± 0.25 mm) based on the distribution of AOD500 measurements by the Reference Grader (Supplementary Figure 2). Measurement agreement between the Reference Grader and the Expert Grader or either algorithm was excellent (ICC range 0.955 to 0.997) and similar for all parameters (Table 1). Measurement agreement for the Novice Grader was lower but still excellent for all parameters (ICC range 0.918 to 0.994). Bland-Altman plots for AOD500 reflected consistent agreement across the entire range of AOD500 measurements for all four (Figure 3). ICCs of measurements from only superior or temporal sectors from one eye per participants showed similar trends (Supplementary Tables 1 and 2) as the primary analysis.

**Table 1.** Human-human and human-machine reproducibility of measurements of scleral spur-based biometric parameters. Intraclass correlation coefficients (ICCs) with 95% confidence intervals comparing measurements from all sectors by the Reference Grader and a second human grader or DL algorithm.

|  | **Expert Grader** | **Novice Grader** | **FPR4** | **TPR95** |
| --- | --- | --- | --- | --- |
| **ACW** | 0.966 (0.962 - 0.970) | 0.943 (0.936 - 0.949) | 0.981 (0.979 - 0.984) | 0.976 (0.973 - 0.978) |
| **LV** | 0.996 (0.995 - 0.996) | 0.994 (0.993 - 0.994) | 0.997 (0.997 - 0.998) | 0.997 (0.996 - 0.997) |
| **AOD500** | 0.961 (0.957 - 0.965) | 0.925 (0.917 - 0.932) | 0.976 (0.974 - 0.979) | 0.973 (0.970 - 0.975) |
| **AOD750** | 0.976 (0.973 - 0.978) | 0.950 (0.945 - 0.955) | 0.981 (0.979 - 0.983) | 0.977 (0.975 - 0.979) |
| **TISA500** | 0.965 (0.961 - 0.968) | 0.918 (0.909 - 0.926) | 0.980 (0.977 - 0.982) | 0.976 (0.974 - 0.979) |
| **TISA750** | 0.974 (0.971 - 0.977) | 0.934 (0.926 - 0.94) | 0.984 (0.982 - 0.986) | 0.981 (0.979 - 0.983) |
| **SSA500** | 0.955 (0.95 - 0.959) | 0.928 (0.92 - 0.935) | 0.970 (0.967 - 0.973) | 0.968 (0.964 - 0.971) |
| **SSA750** | 0.969 (0.966 - 0.972) | 0.945 (0.939 - 0.95) | 0.979 (0.976 - 0.981) | 0.973 (0.971 - 0.976) |

ACW, Anterior Chamber Width. LV, Lens Vault. AOD500/750, Anterior Opening Distance 500/750 μm from the scleral spur. TISA500/750, Trabecular Iris Space Area 500/750 μm from the scleral spur. SSA500/750, Scleral Spur Angle 500/750 μm from the scleral spur.

**Figure 3.**
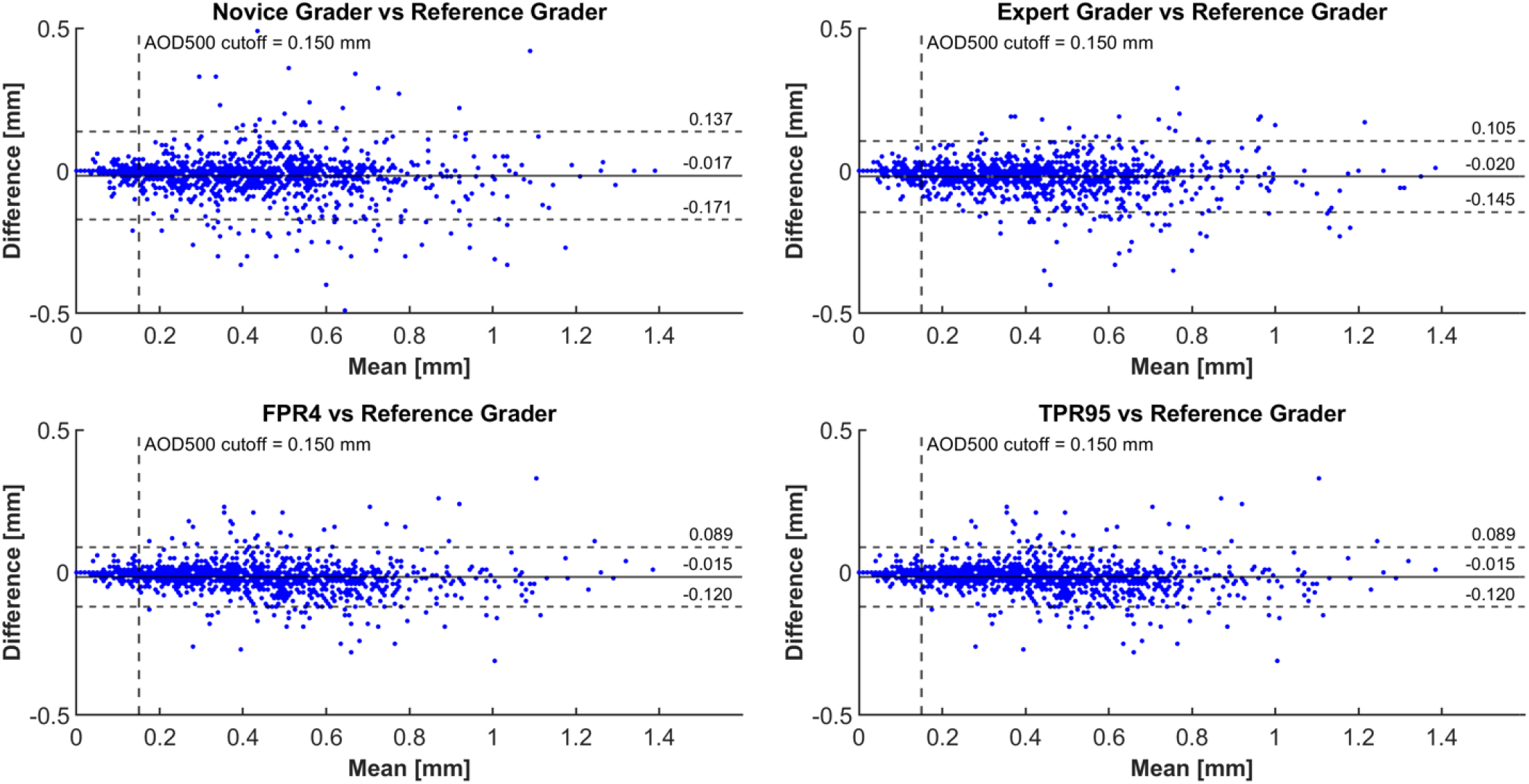
Bland-Altman plots of human-human and human-machine comparisons of AOD500 measurements. Vertical dotted line indicates cutoff (AOD500 < 150 μm) for narrow angles.

Among the 1,504 AS-OCT images graded by the Reference Grader, 198 (13.2%) had narrow angles. Among measurements from these images, ICCs for ACW and LV were similar (ICC range 0.856 to 0.979) whereas ICCs for angle width measurements tended to be lower (ICC range 0.146 to 0.878), primarily due to lower overall variance among the smaller angle width measurements. The differences between Expert and Novice Graders were more pronounced, and the DL algorithms still matched if not exceeded inter-expert agreement (Table 2). Bland-Altman plots for AOD500 demonstrated consistent intra-grader reproducibility, even below the 150 µm threshold for narrow angles (Figure 3).

**Table 2.** Human-human and human-machine reproducibility of measurements of scleral spur-based biometric parameters in narrow angles (AOD500 less than 150 µm). Intraclass correlation coefficients (ICCs) with 95% confidence intervals comparing measurements from all sectors by the Reference Grader and a second human grader or DL algorithm.

|  | **Expert Grader** | **Novice Grader** | **FPR4** | **TPR95** |
| --- | --- | --- | --- | --- |
| **ACW** | 0.931 (0.907 - 0.949) | 0.856 (0.807 - 0.894) | 0.972 (0.958 - 0.981) | 0.959 (0.944 - 0.970) |
| **LV** | 0.965 (0.952 - 0.975) | 0.929 (0.904 - 0.949) | 0.979 (0.970 - 0.986) | 0.973 (0.963 - 0.981) |
| **AOD500** | 0.548 (0.435 - 0.644) | 0.146 (-0.002 - 0.288) | 0.777 (0.699 - 0.836) | 0.746 (0.668 - 0.807) |
| **AOD750** | 0.764 (0.698 - 0.817) | 0.528 (0.416 - 0.623) | 0.861 (0.812 - 0.898) | 0.821 (0.767 - 0.863) |
| **TISA500** | 0.762 (0.682 - 0.824) | 0.267 (0.105 - 0.416) | 0.852 (0.790 - 0.897) | 0.796 (0.721 - 0.852) |
| **TISA750** | 0.792 (0.721 - 0.847) | 0.257 (0.094 - 0.406) | 0.878 (0.826 - 0.916) | 0.824 (0.759 - 0.873) |
| **SSA500** | 0.567 (0.458 - 0.660) | 0.219 (0.073 - 0.355) | 0.782 (0.706 - 0.841) | 0.750 (0.673 - 0.811) |
| **SSA750** | 0.765 (0.699 - 0.818) | 0.574 (0.469 - 0.662) | 0.854 (0.803 - 0.893) | 0.816 (0.761 - 0.859) |

ACW, Anterior Chamber Width. LV, Lens Vault. AOD500/750, Anterior Opening Distance 500/750 μm from the scleral spur. TISA500/750, Trabecular Iris Space Area 500/750 μm from the scleral spur. SSA500/750, Scleral Spur Angle 500/750 μm from the scleral spur.

Rates of FN_ref_ and FP_ref_ differed by grader and algorithm (Figure 4). The Expert and Novice Graders and TPR95 algorithm all had FNR_ref_ < 3.0% and FPR_ref_ > 10.0% whereas the FPR4 algorithm had FNR_ref_ = 12.6% and FPR_ref_ = 9.6%. Compared to the consensus, the FNR_con_ of the FPR4 algorithm was higher than the TPR95 algorithm (12.3% vs 2.7%) whereas the difference in the FPR_con_ was smaller (1.1% vs. 4.1%). On visual inspection of misclassified images by the TPR95 algorithm, many of the images had obvious lid, shadowing, or motion artifacts that make scleral spur detection difficult (Supplementary Figure 3).

**Figure 4.**
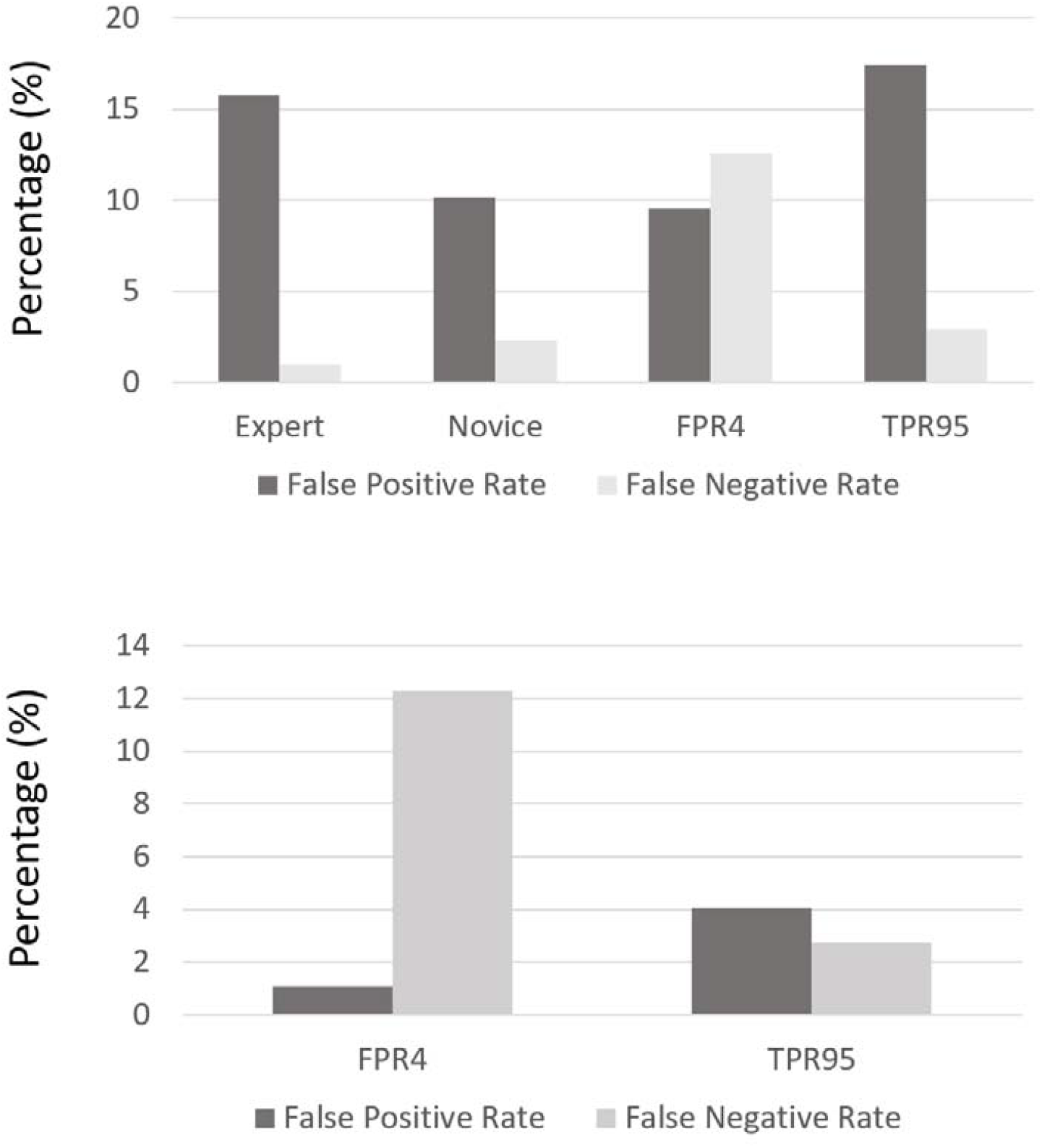
False positive rate (FP) and false negative (FN) rates relative to the Reference Grader (top) and consensus between all three human graders (bottom).

## DISCUSSION

In this study, DL algorithms for the ANTERION OCT system achieved expert-level performance predicting scleral spur locations and measurements of scleral spur-based biometric parameters in a large set of AS-OCT images from an independent patient population and clinical environment. Both the conservative (FPR4) and aggressive (TPR95) algorithms generally approximated the performance of the Expert Grader and exceeded that of the Novice Grader, especially among images with narrow angles. The TPR95 algorithm more closely approximated the FNR and FPR of the human graders, while the FPR4 algorithm made substantially fewer predictions. These findings support the implementation of the TPR95 algorithm for scleral spur detection and automated biometric analysis of ANTERION images, which in turn could greatly enhance the accessibility and utility of quantitative AS-OCT imaging.

Measurements of scleral spur-based biometric parameters are dependent on accurate identification of scleral spur location, which is variable even among experienced graders.^24,25^ Both the FPR4 and TPR95 algorithms produced similar accuracy in predicting scleral spur locations relative to the Reference Grader, with median differences that were smaller than those of the Expert and Novice Graders (<60 μm for both algorithms). This performance is comparable to that of a DL algorithm developed by Xu et al. for the Tomey CASIA SS-1000, in which the mean human-machine scleral spur location difference was 73.08 ± 52.06 μm.^27^ Pham et al. developed a different DL algorithm for the CASIA SS-1000 and plots of human-human and human-machine differences are on a similar scale to those from this study.^26^ These findings suggest that the FPR4 and TPR95 algorithms achieve expert-level performance in scleral spur detection that approximates if not exceeds the agreement between two experienced graders.

Limited access to quantitative measurements of scleral spur-based biometric parameters has hindered the development and implementation of novel clinical methods for evaluating and treating a range of ocular conditions, including PACD, refractive error, and cataract. Our findings suggest that biometric measurements associated with scleral spur predictions by both algorithms are highly correlated with measurements by the Reference Grader and approximate the agreement between two experienced human graders, including in eyes with narrow angles. An automated method that provides access to expert-level measurements of scleral spur-based biometric measurements could help modernize the clinical evaluation and management of patients with PACD. Measurements of AOD and TISA are associated with IOP and anatomical variations in PACD eyes and may predict a higher risk of PACD progression or poor angle widening after LPI.^8,15,16^ In addition, automated measurements of ACW and LV could be beneficial for IOL selection: ACW is helpful in sizing anterior chamber and phakic IOLs, and there is evidence that LV could play an important role in determining effective lens position and calculating IOL power.^18–23^

Our results demonstrate that rates of scleral spur detection are highly variable under real-world conditions without eyelid retraction during imaging, even among experienced graders. This point, which has not been previously studied, suggests there is differing confidence among graders when deciding whether to mark a scleral spur. Based on number of scleral spurs marked, the Reference Grader appeared the most conservative and the Expert Grader the most aggressive among human graders. The TPR95 algorithm approximated the FNR and FPR of the Expert Grader (1.0% and 15.8% vs. 2.9% and 17.4%). While the more conservative FPR4 algorithm had a lower FPR compared to the TPR95 algorithm (9.6% vs. 17.4%), this came at the expense of a higher FNR (12.6% vs. 2.9%). Despite the greater number of scleral spurs identified by the TPR95 algorithm, measurement agreement between the Reference Grader and both algorithms were similar. In a busy clinical environment, the higher TPR of the TPR95 algorithm is likely of greater utility than the lower FPR of the FPR4 algorithm as it is more convenient to ignore a questionably marked scleral spur than to manually mark a more obvious one.

Our study has several strengths compared to prior studies on automated scleral spur detection.^26,27^ First, the DL algorithms maintained expert-level performance in a diverse patient cohort and real-world clinical environment that was completely independent from the cohort and environment in which the algorithm was developed. These findings support the generalizability and widespread implementation of DL algorithms in diverse practice settings, while prior studies that used smaller and more homogenous cohorts do not.^26,27^ Second, images with eyelid or other imaging artifacts were not omitted from in the validation dataset. This approach allowed us to assess variability in human grader and algorithm confidence in scleral spur detection and evaluate its effect on detection rates and measurement agreement. It also avoids introducing biases associated with analyzing only a subset of images and applying arbitrary definitions of image quality that may be difficult in real-world practice environments. Third, all images were graded by a novice grader in addition to a second expert grader, which allowed us to determine that there is a benefit to using DL algorithms over a trained but inexperienced grader.

Our study also has several limitations. First, the Reference Grader was relatively conservative and marked fewer images than the other human graders and TPR95 algorithm. Second, while the overall number of images analyzed was large, there were fewer images and subsequently wider ICC confidence intervals in sub-analyses that accounted for angle width and intra-and inter-eye correlations. In the future, a larger cohort would be beneficial for a more detailed study of measurements from individual sectors of the eye. Third, the described algorithms are only available for images acquired on the ANTERION OCT system, and their expert-level performance would likely not generalize to images acquired on other AS-OCT devices.

In conclusion, DL algorithms provide expert-level scleral spur detection and biometric analysis in a large set of AS-OCT images from a diverse clinical cohort. There appears to be a benefit to using the TPR95 algorithm compared to grading by a novice in terms of the number of scleral spurs identified and the accuracy of biometric measurements. This study supports the implementation of the TPR95 algorithm in diverse patient populations and real-world practice settings, which could help expand the clinical utility of AS-OCT imaging and modernize the care of ocular conditions dependent on accurate anterior segment biometry.

## Data Availability

Data is not available

## ACKNOWLEDGEMENTS

This work was supported by grant K23 EY029763 from the National Eye Institute, National Institute of Health, Bethesda, Maryland and an unrestricted grant to the Department of Ophthalmology from Research to Prevent Blindness, New York, NY.

## DISCLOSURES

K.B., G.A., A.A.P., B.B., and X.X. have no relevant financial disclosures. B.Y.X. and A.S.H. receive research support from Heidelberg Engineering. M.S. is employed by Heidelberg Engineering.

## TABLE AND FIGURE CAPTIONS

**Supplementary Figure 1**. Receiver operating characteristic (ROC) curve for scleral spur prediction by the DL algorithm in Heidelberg Engineering’s internal test dataset.

**Supplementary Figure 2**. Distribution of AOD500 as measured by the Reference Grader.

**Supplementary Table 1**. Intraclass correlation coefficients (ICCs) with 95% confidence intervals comparing measurements from the superior sector by the Reference Grader and a second human grader or DL algorithm.

**Supplementary Table 2**. Intraclass correlation coefficients (ICCs) with 95% confidence intervals comparing measurements from the temporal sector by the Reference Grader and a second human grader or DL algorithm.

**Supplementary Figure 3**. Representative cropped images of false positives (FP; top) and false negatives (FN; bottom) by the TPR95 algorithm based on the consensus between all three human graders. Red dots in FP images indicate predicted scleral spur location by FPR95 algorithm. Red dots in FN images indicate marked scleral spur location by the Reference Grader.

## Notes

### Author Declarations

Institutional Review Board of the University of Southern California (USC) gave ethical approval for this work

