## Supplementary Figure 1 for "Automated Expert-level Scleral Spur Detection and Quantitative Biometric Analysis on the ANTERION Anterior Segment OCT System"

**Supplementary Figure 1.** Receiver operating characteristic (ROC) curve for scleral spur prediction by the DL algorithm in Heidelberg Engineering’s internal test dataset.

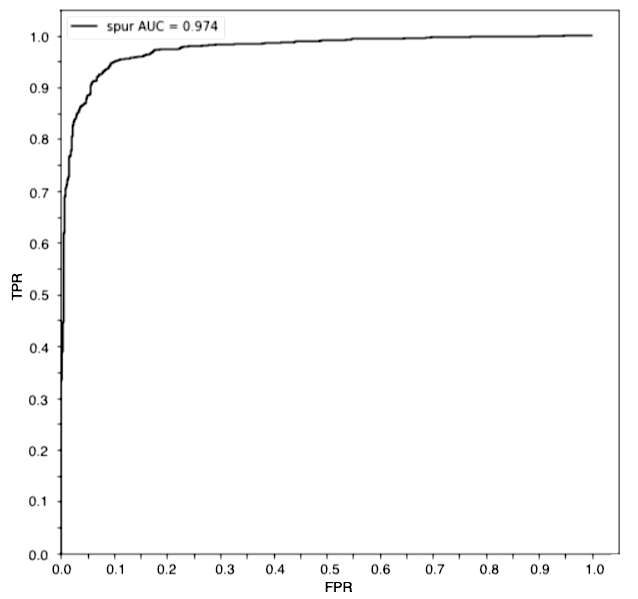

TPR, True Positive Rate. FPR, False Positive Rate. AUC, Area Under Curve.
