## Supplementary Figure 2 for "Automated Expert-level Scleral Spur Detection and Quantitative Biometric Analysis on the ANTERION Anterior Segment OCT System"

**Supplementary Figure 2.** Distribution of AOD500 as measured by the Reference Grader.

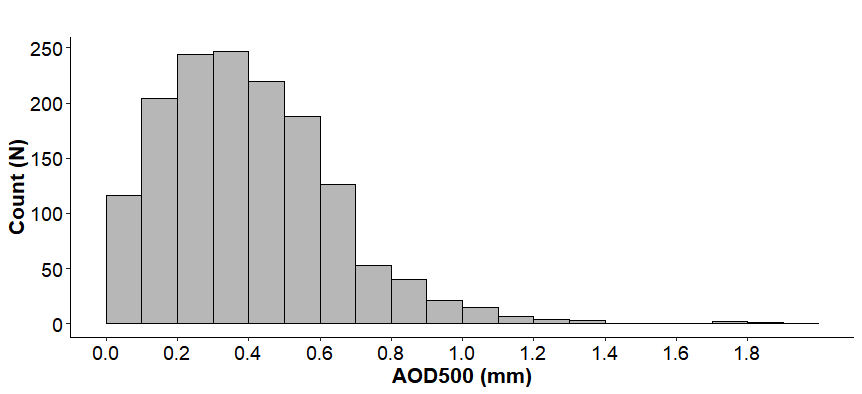

AOD500, Anterior Opening Distance 500 μm from the scleral spur.
