## Supplementary Table 1 for "Automated Expert-level Scleral Spur Detection and Quantitative Biometric Analysis on the ANTERION Anterior Segment OCT System"

**Supplementary Table 1.** Intraclass correlation coefficients (ICCs) with 95% confidence intervals comparing measurements from the superior sector by the Reference Grader and a second human grader or DL algorithm.

|  | **Expert Grader** | **Novice Grader** | **FPR4** | **TPR95** |
| --- | --- | --- | --- | --- |
| **ACW** | 0.891 (0.743 - 0.956) | 0.531 (0.095 - 0.799) | 0.970 (0.903 - 0.991) | 0.956 (0.889 - 0.983) |
| **LV** | 0.996 (0.988 - 0.998) | 0.990 (0.972 - 0.997) | 0.998 (0.993 - 0.999) | 0.996 (0.990 - 0.999) |
| **AOD500** | 0.981 (0.961 - 0.991) | 0.907 (0.808 - 0.956) | 0.989 (0.976 - 0.995) | 0.986 (0.971 - 0.993) |
| **AOD750** | 0.970 (0.939 - 0.986) | 0.950 (0.897 - 0.977) | 0.994 (0.987 - 0.997) | 0.975 (0.950 - 0.988) |
| **TISA500** | 0.957 (0.908 - 0.980) | 0.782 (0.557 - 0.901) | 0.976 (0.944 - 0.990) | 0.946 (0.882 - 0.976) |
| **TISA750** | 0.97 (0.934 - 0.986) | 0.871 (0.723 - 0.943) | 0.985 (0.965 - 0.994) | 0.962 (0.916 - 0.983) |
| **SSA500** | 0.976 (0.95 - 0.989) | 0.925 (0.843 - 0.965) | 0.983 (0.963 - 0.993) | 0.978 (0.953 - 0.989) |
| **SSA750** | 0.961 (0.921 - 0.981) | 0.946 (0.887 - 0.974) | 0.99 (0.979 - 0.996) | 0.97 (0.939 - 0.986) |

ACW, Anterior Chamber Width. LV, Lens Vault. AOD500/750, Anterior Opening Distance 500/750 μm from the scleral spur. TISA500/750, Trabecular Iris Space Area 500/750 μm from the scleral spur. SSA500/750, Scleral Spur Angle 500/750 μm from the scleral spur.
