## Supplementary Table 2 for "Automated Expert-level Scleral Spur Detection and Quantitative Biometric Analysis on the ANTERION Anterior Segment OCT System"

**Supplementary Table 2.** Intraclass correlation coefficients (ICCs) with 95% confidence intervals comparing measurements from the temporal sector by the Reference Grader and a second human grader or DL algorithm.

|  | **Expert Grader** | **Novice Grader** | **FPR4** | **TPR95** |
| --- | --- | --- | --- | --- |
| **ACW** | 0.955 (0.942 - 0.966) | 0.927 (0.905 - 0.943) | 0.980 (0.973 - 0.985) | 0.978 (0.971 - 0.983) |
| **LV** | 0.996 (0.994 - 0.997) | 0.994 (0.993 - 0.996) | 0.996 (0.995 - 0.997) | 0.996 (0.995 - 0.997) |
| **AOD500** | 0.965 (0.955 - 0.973) | 0.950 (0.936 - 0.962) | 0.979 (0.972 - 0.984) | 0.979 (0.972 - 0.984) |
| **AOD750** | 0.977 (0.970 - 0.982) | 0.969 (0.960 - 0.977) | 0.984 (0.980 - 0.988) | 0.984 (0.979 - 0.988) |
| **TISA500** | 0.974 (0.965 - 0.981) | 0.977 (0.969 - 0.983) | 0.986 (0.981 - 0.990) | 0.986 (0.981 - 0.990) |
| **TISA750** | 0.981 (0.975 - 0.986) | 0.978 (0.970 - 0.984) | 0.988 (0.983 - 0.991) | 0.988 (0.983 - 0.991) |
| **SSA500** | 0.944 (0.927 - 0.957) | 0.910 (0.884 - 0.931) | 0.971 (0.962 - 0.978) | 0.970 (0.961 - 0.977) |
| **SSA750** | 0.964 (0.953 - 0.972) | 0.942 (0.925 - 0.956) | 0.983 (0.978 - 0.987) | 0.982 (0.976 - 0.986) |

ACW, Anterior Chamber Width. LV, Lens Vault. AOD500/750, Anterior Opening Distance 500/750 μm from the scleral spur. TISA500/750, Trabecular Iris Space Area 500/750 μm from the scleral spur. SSA500/750, Scleral Spur Angle 500/750 μm from the scleral spur.
