## Supplementary Figure 3 for "Automated Expert-level Scleral Spur Detection and Quantitative Biometric Analysis on the ANTERION Anterior Segment OCT System"

**Supplementary Figure 3.** Representative cropped images of false positives (FP; top) and false negatives (FN; bottom) by the TPR95 algorithm based on the consensus between all three human graders. Red dots in FP images indicate predicted scleral spur location by FPR95 algorithm. Blue dots in FN images indicate scleral spur location marked by the Reference Grader.

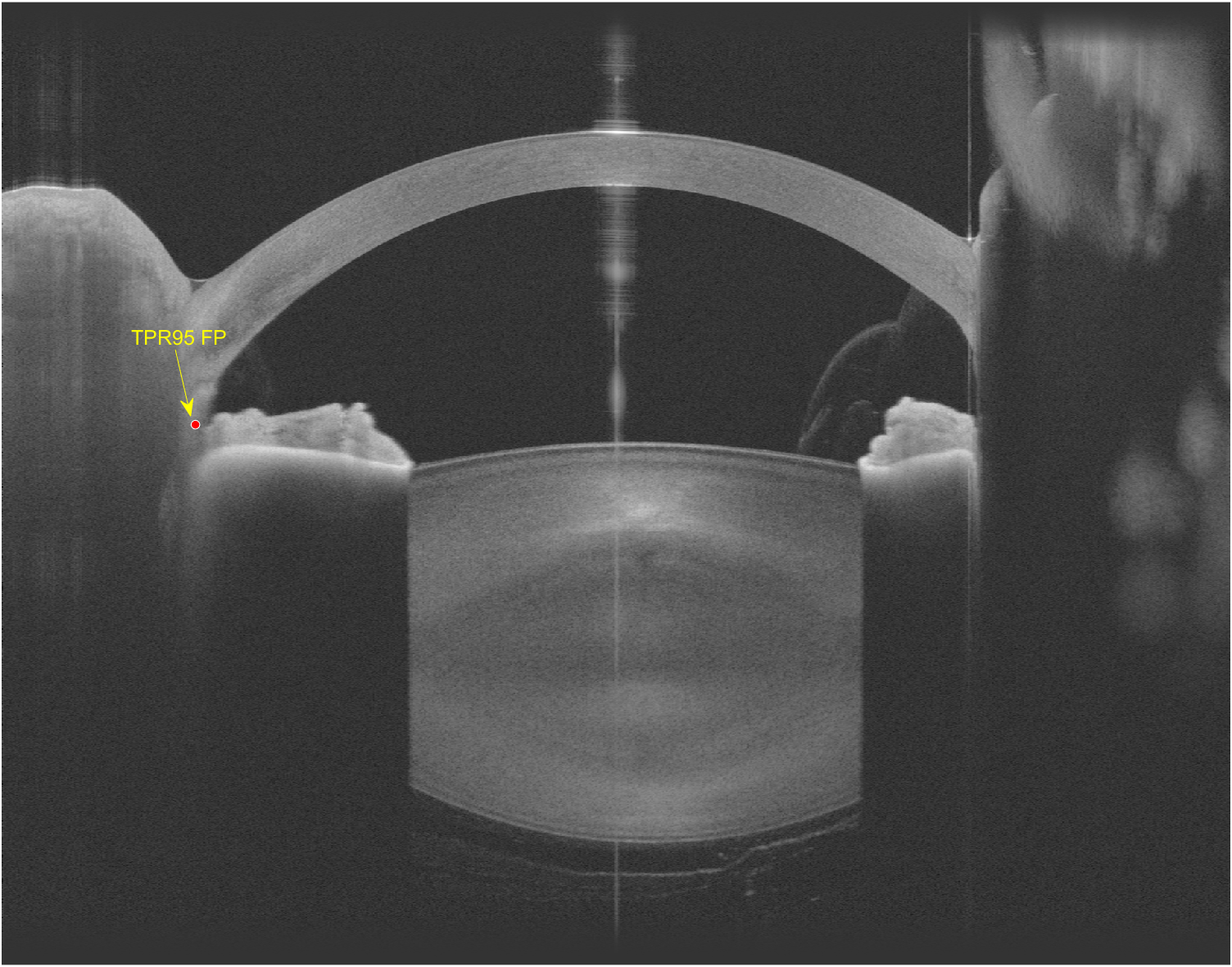

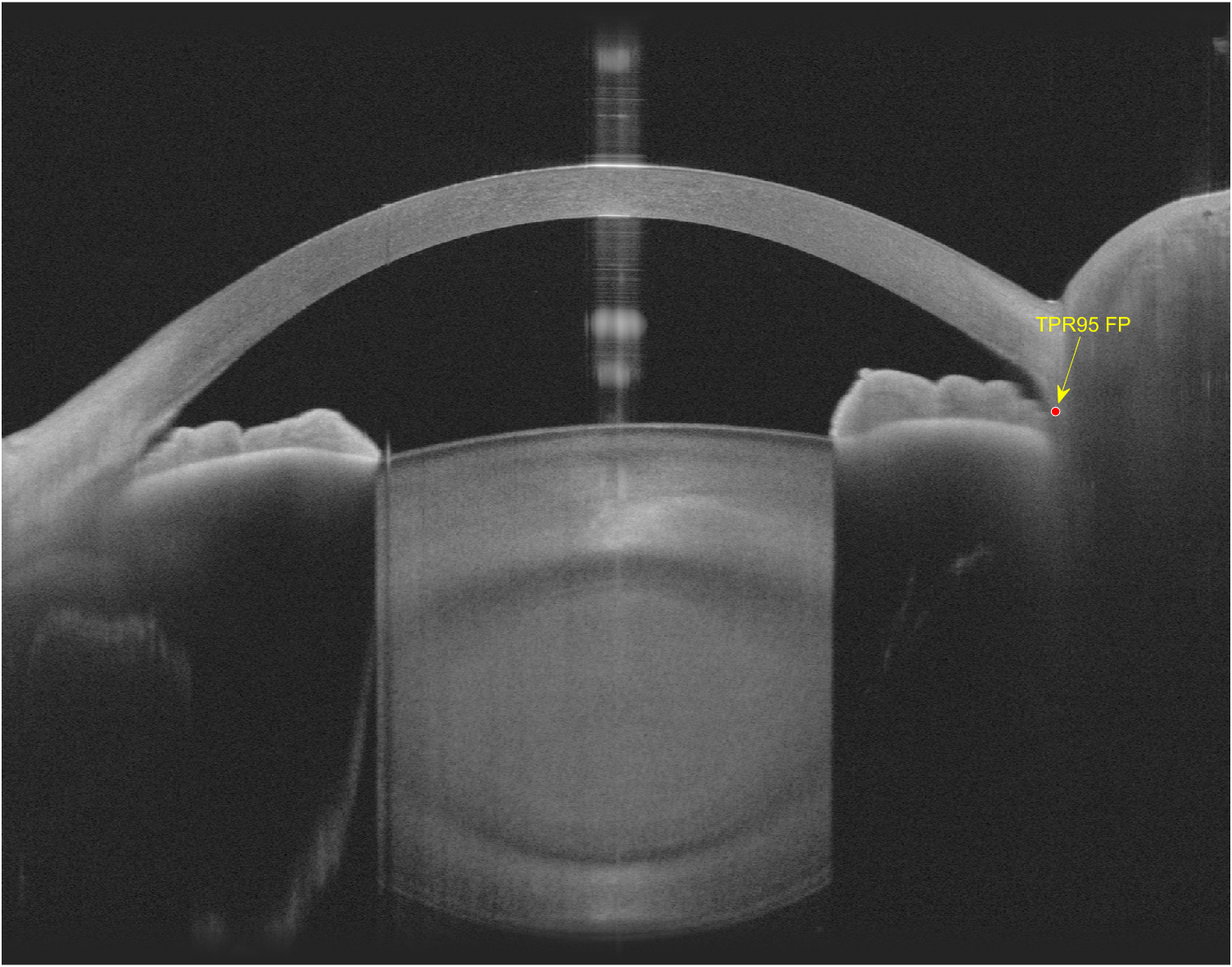

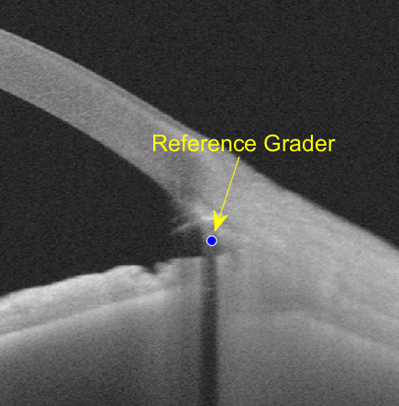

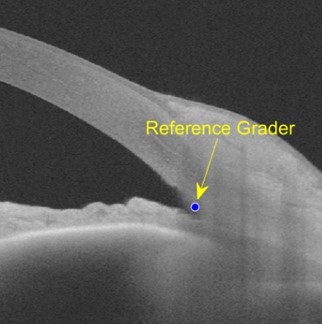
